# Estimated Head Motion Contributes to Case-Control Magnetic Resonance Imaging Morphometry Differences in Schizophrenia

**DOI:** 10.64898/2026.03.04.26347600

**Authors:** Roberta Passiatore, Nicola Sambuco, Giuseppe Stolfa, Linda A. Antonucci, Alessandro Bertolino, Giuseppe Blasi, Leonardo Fazio, Aaron L. Goldman, Luigi Grassi, Daniela Grasso, Annchen R. Knodt, Antonella Lupo, Ciro Mazza, Alessio M. Monteleone, Antonio Rampino, William S. Ulrich, Ethan T. Whitman, Ahmad R. Hariri, Daniel R. Weinberger, the Apulian Network on Risk for Psychosis, Giulio Pergola

## Abstract

In-scanner head motion is a recognized source of bias in structural magnetic resonance imaging (sMRI), yet it remains under-addressed in psychiatric neuroimaging where structural difference in patient populations are considered foundational. We examined motion-related bias in grey matter volume estimates across eight independent cohorts comprising 9,664 individuals, including 8,979 neurotypical controls (NC), 497 patients with schizophrenia (SCZ), and 188 patients with bipolar disorder (BD). Motion estimates were derived from multiple fMRI scans acquired within the same scanning session and summarized using principal component analysis. In NC, motion accounted for 1–6% of regional grey matter variance, a magnitude comparable to reported psychiatric case–control effect sizes. Adjusting for motion attenuated SCZ–NC group differences, reducing effect sizes in 85% of brain regions and yielding 5% fewer significant ROIs (pFDR<0.05). In BD, motion correction reduced effect sizes in 97% of regions, with a 24% reduction in significant ROIs. Cross-diagnostic spatial patterns were significantly correlated (r=0.63, p=3×10⁻¹³), explaining a sizable portion of SCZ-BD commonalities. Critically, a falsification analysis in UK Biobank (N=5,123) showed that stratifying NC by motion alone produced grey matter differences accounting for 45–62% of SCZ case-control effect magnitude, underscoring how difficult it is to interpret SCZ-like morphometric differences as tissue properties rather than as motion-driven patterns. These findings urge caution in interpretations of sMRIdifferences in patient-control comparisons and use of systematic fMRI based motion control as standard practice in sMRI analyses.

## 1. Introduction

Brain Magnetic Resonance Imaging (MRI) is the primary modality for *in vivo* non-invasive neuroanatomical assessment in humans. Advances in automated brain segmentation and morphometric approaches have enabled large-scale investigations of neuroanatomical variation and spurred interest in leveraging MRI-derived indicators for diagnosis and risk prediction in neuropsychiatric disorders ^1–3^. However, MRI does not directly quantify biological tissue properties. Instead, it reflects the interaction between acquisition parameters and tissue-dependent physical and chemical properties. Therefore, regional differences represent composite signal variations rather than direct measures of cellular composition or cytoarchitecture. Consequently, group differences in structural MRI (sMRI) between patients without evident neuropathology and neurotypical controls (NC) do not necessarily reflect the actual variations in brain tissue ^4,5^.

In-scanner head motion is a well-documented source of bias in sMRI estimates ^6–8^. Psychiatric, neurologic, and developmental populations exhibit greater motion than controls, leading to differential exclusion rates during quality control procedures ^9,10^. Although prospective motion-tracking and correction approaches can mitigate motion artifacts during scanning ^11^, retrospective solutions commonly applied to existing datasets, particularly in the context of large consortia drawing on historical data collection, remain insufficient to remove residual motion effects on cortical morphometry ^12^.

Previous studies have shown that motion induces systematic variation in grey matter estimates in NC ^13^ and in clinical populations, with framewise displacement correlating with reduced morphometric estimates in schizophrenia (SCZ) and bipolar disorder (BD) ^14,15^. Yet, despite broad acknowledgment of this issue, no large-scale study has directly tested the extent to which motion proxies inflate case-control morphometric differences in psychiatric populations.

Here, we analyzed data of 9,664 individuals drawn from eight independent cohorts. We estimated the proportion of variance in grey matter volumes explained by in-scanner head motion by quantifying motion from fMRI scans collected only a few minutes apart (Table 1). Analyses included 8,979 NC scans, of which 5,123 were drawn from the UK Biobank. One to three functional scans were available per participant, encompassing one resting-state session (RS) and, when available, task-based sessions including a working memory task (WM) and explicit or implicit emotional face processing tasks (FE and FI) ^16–19^. We reasoned that if volumetric estimates are significantly explained by in-scanner motion in NC, then reported case-control morphometric differences are likely confounded by motion-related effects rather than necessarily reflecting disease-specific neuroanatomy, particularly if psychiatric populations exhibit systematically greater head motion ^20^. For instance, if greater head motion is associated with lower grey matter estimates, high-motion NC would spuriously appear “patient-like”, whereas higher motion in patients would overestimate disease-related atrophy.

**Table 1.** Demographic characteristics and MRI acquisition details for each cohort in the study. *Abbreviations.* SCZ: Patients with schizophrenia; BD: Patients with bipolar disorder; NC: Neurotypical controls; SD: Standard deviation; Vx: voxel dimension; FOV: field of view; Vol: number of volumes; TR: Repetition time in ms; TE: Time to echo in ms.

| Cohort<br>(MRI<br>Scanner) | MRI<br>Sequenc<br>e Type | MRI Parameters | Sample |  |  |  |  |  |
| --- | --- | --- | --- | --- | --- | --- | --- | --- |
|  |  |  | SCZ |  | BD |  | NC |  |
|  |  |  | N (F) | Age -<br>Mean<br>(SD) | N (F) | Age -<br>Mean<br>(SD) | N (F) | Age -<br>Mean<br>(SD) |
| **LIBD**<br>(General<br>Electric<br>Signa<br>3T) | Structura<br>l MRI | MPRAGE; Vx:<br>1×1×1.3 mm;<br>FOV/slice: 256/124 | - |  |  |  |  |  |
|  | Working<br>memory<br>fMRI | GE-EPI;<br>TR/TE:2000/28;<br>Vol:120;<br>Vx:3.75×3.75×5<br>mm;<br>FOV/slice:240/20 | 134<br>(43) | 32.2<br>(10.4) | - | - | 485<br>(277) | 32.7<br>(10.5) |
|  | Resting<br>state<br>fMRI | GE-EPI; TR/TE:<br>1600/26; Vol:300;<br>Vx:4×4×5 mm;<br>FOV/slice:256/28 | 115<br>(39) | 31.2<br>(9.9) | - | - | 250<br>(156) | 30.6<br>(9.8) |
|  | Implicit<br>emotion<br>recogniti<br>on task<br>fMRI | GE-EPI;<br>TR/TE:2000/28;<br>Vol:144;<br>Vx:3.75×3.75×5<br>mm;<br>FOV/slice:240/20 | 102<br>(36) | 31.0<br>(9.9) | - | - | 390<br>(225) | 32.8<br>(10.5) |
| **UNIBA1**<br>(General<br>Electric | Structura<br>l MRI | MPRAGE; Vx:<br>1×1×1.3 mm;<br>FOV/slice: 256/124 | - |  |  |  |  |  |
| Signa 3T) | Working memory fMRI | GE-EPI; TR/TE:2000/28; Vol:120; Vx:3.75×3.75×5 mm; FOV/slice:240/20 | 64 (21) | 32.9 (8.5) | - | - | 405 (221) | 26.5 (7.7) |
|  | Explicit emotion recognition task fMRI | GE-EPI; TR/TE:2000/28; Vol:180; Vx:3.75×3.75×5 mm; FOV/slice:240/20 | 55 (18) | 33.2 (9.1) | - | - | 342 (173) | 26.8 (7.9) |
|  | Implicit emotion recognition task fMRI | GE-EPI; TR/TE:2000/28; Vol:180; Vx:3.75×3.75×5 mm; FOV/slice:240/20 | 37 (13) | 34.8 (9.3) | - | - | 244 (124) | 26.5 (7.8) |
| UNIBA2 (Philips Ingenia 3T) | Structural MRI | T1-FFE; Vx: 1×1×1 mm; FOV/slice: 256/180 | - |  |  |  |  |  |
|  | Working memory fMRI | EPI; TR/TE:3000/38; Vol:120; Vx:3×3×3.6 mm; FOV/slice:240/38 | 41 (8) | 29.8 (8.5) | 39 (16) | 35.0 (12.7) | 196 (103) | 27.0 (7.1) |
|  | Resting state fMRI | EPI; TR/TE:3000/38; Vol:240; Vx:3×3×3.6 mm; FOV/slice:240/38 | 46 (12) | 29.9 (8.5) | 48 (19) | 35.0 (12.7) | 235 (131) | 27.0 (7.3) |
|  | Implicit emotion recognition task fMRI | EPI; TR/TE:3000/38; Vol:212; Vx:3×3×3.6 mm; FOV/slice:240/38 | 31 (10) | 30.4 (9.6) | 40 (14) | 34.9 (12.3) | 152 (89) | 29.0 (7.2) |
| HCP (Siemens Connecto | Structural MRI | MPRAGE; Vx: 0.7×0.7×0.7 mm; FOV/slice: 224/256 | - |  |  |  |  |  |
| me Skyra<br>3T) | Working<br>memory<br>fMRI | GE-EPI;<br>TR/TE:720/33.1;<br>Vol:405; Vx:2×2×2<br>mm;<br>FOV/slice:208/72 | - | - | - | - | 1,031<br>(560) | 27.0<br>(3.5) |
|  | Resting<br>state<br>fMRI | GE-EPI;<br>TR/TE:720/33.1;<br>Vol:1200; Vx:2×2×2<br>mm;<br>FOV/slice:208/72 | - | - | - | - | 1,031<br>(560) | 27.0<br>(3.5) |
|  | Implicit<br>emotion<br>recogniti<br>on task<br>fMRI | GE-EPI;<br>TR/TE:720/33.1;<br>Vol:176; Vx:2×2×2<br>mm;<br>FOV/slice:208/72 | - | - | - | - | 1,031<br>(560) | 27.0<br>(3.5) |
| **CNP**<br>(Two<br>identical<br>Siemens<br>Trio 3T) | Structura<br>l MRI | MPRAGE;<br>Vx:0.98×0.98×1<br>mm; FOV/slice:<br>250/176 | - |  |  |  |  |  |
|  | Working<br>memory<br>fMRI | GE-EPI;<br>TR/TE:2000/30;<br>Vol:295; Vx:3×3×4<br>mm;<br>FOV/slice:192/34 | 44<br>(11) | 36.3<br>(9.1) | 48<br>(20) | 35.6<br>(8.9) | 117<br>(54) | 31.3<br>(8.6) |
|  | Resting<br>state<br>fMRI | GE-EPI;<br>TR/TE:2000/30;<br>Vol:156; Vx:3×3×4<br>mm;<br>FOV/slice:192/34 | 44<br>(11) | 36.3<br>(9.1) | 48<br>(20) | 35.6<br>(8.9) | 117<br>(54) | 31.3<br>(8.6) |
| **DNS**<br>(Two<br>identical<br>GE<br>MR750<br>3T) | Structura<br>l MRI | FSPGR;<br>Vx:0.94×0.94×1<br>mm;<br>FOV/slice:240/162 | - |  |  |  |  |  |
|  | Working<br>memory<br>fMRI | GE-EPI;<br>TR/TE:2000/30;<br>Vol:354; Vx:<br>3.75×3.75×4 mm;<br>FOV/slice:240/34 | - | - | - | - | 298<br>(171) | 20.3<br>(1.3) |
|  | Resting state fMRI | GE-EPI; TR/TE:2000/30; Vol:128; Vx: 3.75×3.75×4 mm; FOV/slice:240/34 | - | - | - | - | 977 (561) | 20.2 (1.3) |
|  | Implicit emotion recognition task fMRI | GE-EPI; TR/TE:2000/30; Vol:195; Vx: 3.75×3.75×4 mm; FOV/slice:240/34 | - | - | - | - | 1,284 (738) | 20.2 (1.3) |
| **BSNIP**<br>(GE: Signa, Signa HDxt; Philips: Achieva; Siemens: Allegra, Trio) | Structural MRI (range) | MPRAGE – IR-SPGR; Vx:1×1×1.2 mm; FOV/slice: 240-260/160-170 | - |  |  |  |  |  |
|  | Resting state fMRI (range) | GE-EPI; TR/TE: 1500-3000/22-30; Vol:100-200; Vx:3.4×3.4×3-5 mm; FOV/slice: 218×218×87-150/29-36 | 141 (45) | 34.4 (12.3) | 88 (57) | 36.9 (12.0) | 169 (104) | 39.2 (12.8) |
| **UK Biobank**<br>(Siemens Skyra 3T) | Structural MRI | MPRAGE; Vx: 1×1×1 mm; FOV/slice: 256/208 | - |  |  |  |  |  |
|  | Resting state fMRI | Multiband-EPI; TR/TE:735/39; Vol:490; Vx: 2.4×2.4×2.4 mm; FOV/slice:210/60 | - | - | - | - | 5,123 (2,563) | 52.7 (7.3) |
|  | Implicit emotion recognition task fMRI | Multiband-EPI; TR/TE:735/39; Vol:332; Vx: 2.4×2.4×2.4 mm; FOV/slice:210/60 | - | - | - | - | 4,802 (2,434) | 52.5 (7.3) |

To test these hypotheses, we focused primarily on SCZ, a disorder characterized by large and spatially widespread volumetric differences reported by ∼500 peer-reviewed studies involving more than 38,000 patients and NC ^21^. Rather surprisingly, reported SCZ-NC effect sizes approach those observed in neurodegenerative disorders ^22,23^, and exceed differences reported across other major psychiatric and neurodevelopmental conditions, including BD, major depressive disorder, and autism spectrum disorder ^24^, despite limited confirmation in postmortem studies ^25–27^. Thus, SCZ is an ideal benchmark for testing to what extent motion may inflate morphometric differences. We compared grey matter volumes between NC and SCZ using (i) a standard model without motion-related covariates, and (ii) a motion-adjusted model including principal components (PCs) derived from fMRI-based motion estimates. Given the systematic association between diagnosis and motion ^14^, we expected that accounting for motion would attenuate SCZ-NC group differences.

To assess diagnostic specificity, we conducted a secondary validation analysis in an independent sample of 188 patients with BD type I and II. Based on prior evidence that sMRI differences are largely nonspecific across psychiatric diagnoses ^24^, we expected a similar attenuation of group differences after motion adjustment, along with overlapping spatial patterns of motion susceptibility between SCZ and BD that may explain the purported grey matter deficit in a psychosis continuum framework.

Finally, a large sample of psychiatrically and neurologically unaffected participants drawn from the UK Biobank (N=5,123) enabled a diagnosis-free falsification analysis. Within this cohort, we compared NC individuals with high versus low head motion (top vs. bottom deciles of the first motion-derived principal component, PC1) using the identical analytic pipeline applied to case-control comparisons. This within-sample design directly tested whether motion alone could generate morphometric differences resembling those reported in patient populations.

Together, this multi-cohort framework provides a direct assessment of the extent to which in-scanner head motion contributes to, and potentially inflates, reported case–control morphometric differences in psychiatric neuroimaging, with broad implications for the interpretation of neuroanatomical differences across psychiatric disorders.

## 2. Results

### 2.1 Motion-related estimates from fMRI scans

To obtain a low-dimensional summary of in-scanner head motion, we derived motion estimates from all available fMRI scans across the eight cohorts (Table 1) and reduced them using principal component analysis (PCA). The first five PCs (PC1–PC5) captured the dominant axes of motion variability and together explained >80% of variance in the motion feature set (Figure 1). Accordingly, PC1–PC5 were carried forward as covariates to account for motion-related variability in subsequent analyses.

**Figure 1.**
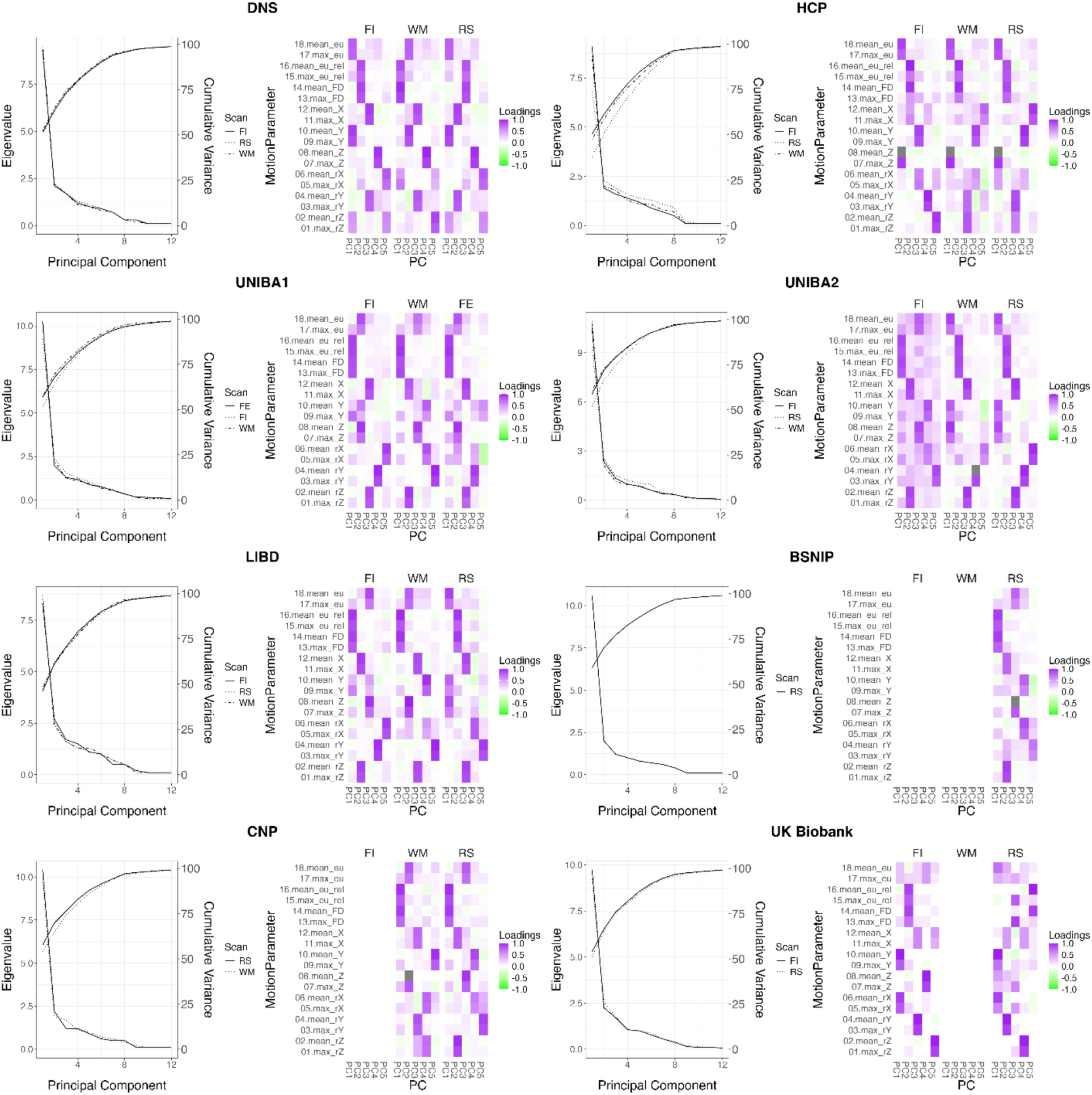
Left. The figure illustrates the eigenvalues and cumulative variance explained across principal components from the eight cohorts used in the current study. Right. The loadings of the first five principal components follow oblique rotation (oblimin) in each of the eight cohorts. Abbreviations. FE: Faces – Explicit; FI: Faces – Implicit; WM: Working memory; RS: Resting-State

Motion PCs showed high consistency across cohorts and fMRI scan types (Figure S1). In the DNS cohort, for instance, PC1 was highly correlated between the RS and FI scans (r = 0.99, pFDR = 2 × 10^−11^), indicating robust reliability across acquisition types. Similarly, in clinical cohorts such as UNIBA2 and LIBD, corresponding PCs derived from the FI and RS scans also showed strong correspondence (PC2: r = 0.969, pFDR = 6 ×10^−10^ and r = 0.960, pFDR = 3 × 10^−10^, respectively). Across cohorts and scan types, cross-site and cross-scan correlations exceeded r = 0.9 and survived FDR correction (Figure S1).

Within cohorts, PC scores were also significantly correlated across different fMRI scans, such that individuals with higher scores in one scan tended to exhibit higher motion across other scans (e.g., UNIBA1: WM-FE: r = 0.45, p = 2 ×10^−15^; WM-FI: r = 0.52, p = 6.68 ×10^−15^; Table S1), indicating that motion represents a replicable individual characteristic rather than only scan-specific noise.

Motion PCs further differed consistently by diagnostic group, with patients with SCZ and BD showing significantly higher PC scores compared to NC (all p < 0.05; Tables S2-S3). Notably, these group differences were most pronounced for the first three PCs, which together accounted for ∼60% of the total variance captured by the motion-PCs (Figure 1). Distributions of all PCs across cohorts, groups, and fMRI scan types are shown in Figure S2.

### 2.2 Motion effects on gray matter volume estimates in NC

To quantify the impact of in-scanner head motion on structural MRI estimates in NC, we examined the linear associations of motion-derived PCs and regional grey matter volumes in 100 cortical ROIs from the Schaefer-Yeo atlas ^28^ and eight bilateral subcortical ROIs from the Hammersmith atlas ^29,30^. Motion-related effects were highly consistent across fMRI scan types, with significant associations observed in 99%, 98%, and 99% of cortical and subcortical ROIs when motion parameters derived from RS, WM, and FI sessions, respectively. P values were summarized across cohorts through Fisher’s method ^31^ and corrected for multiple comparisons through FDR (pFDR<0.05).

Across regions and cohorts, the 5 motion-PCs accounted for 1 to 6% of the variance in grey matter volume estimates, as quantified by R² and low between-ROI heterogeneity (I² <10) ^32^. The highest value was observed in the thalamus in UK Biobank. Motion-related effects were most prominent in the insular cortex, temporal parietal cortices, lateral and ventral prefrontal cortices, as well as in the thalamus, putamen, and lateral ventricles. Notably, the spatial pattern and magnitude of motion effects were comparable regardless of the fMRI scan used to estimate motion (RS, WM, or FI). Aggregated results across cohorts are shown in Figure 2, while cohort-specific results are shown in Figure S3.

**Figure 2.**
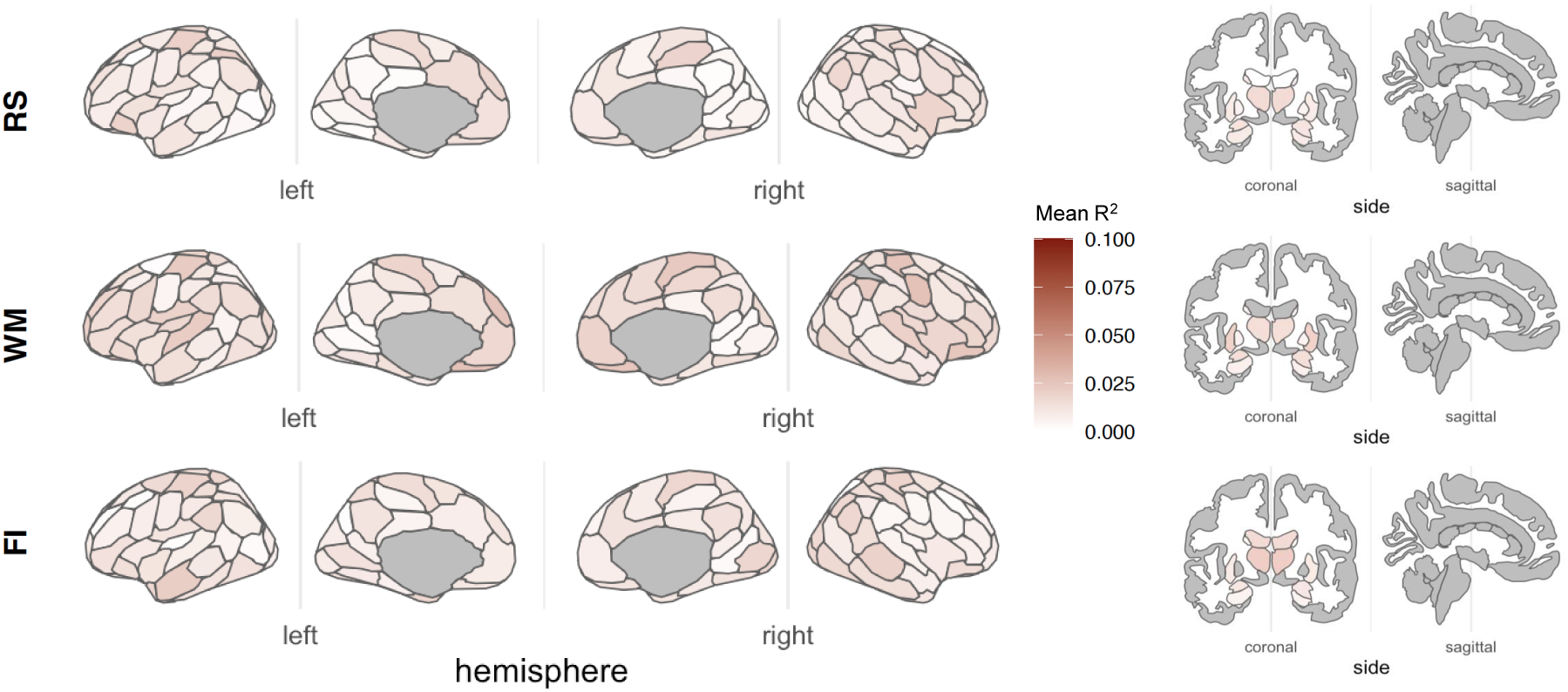
Mean R-squared is depicted, averaging the effects reported across cohorts in cortical and subcortical ROIs. Results are shown at pFDR<0.05. Abbreviations: WM=working memory, RS=resting state, FI=implicit emotional faces recognition.

### 2.3 Motion affects group differences comparing SCZ vs. NC

We assessed group differences between SCZ and NC using standard or motion-corrected analyses, aggregating results across cohorts and fMRI scan types via a multi-level meta-analysis accounting for repeated measures within the same cohort (mixed effects: cohort and session; see Methods). Across ROIs, patients with SCZ showed reduced grey matter volume in the insular cortex, temporoparietal regions, lateral and ventral prefrontal cortices, hippocampus, amygdala, and thalamus, alongside increased volumes in the pallidum and, to a lesser extent, in the lateral ventricles (pFDR < 0.05; Figure 3a), consistent with prior work ^29,33–37^.

**Figure 3.**
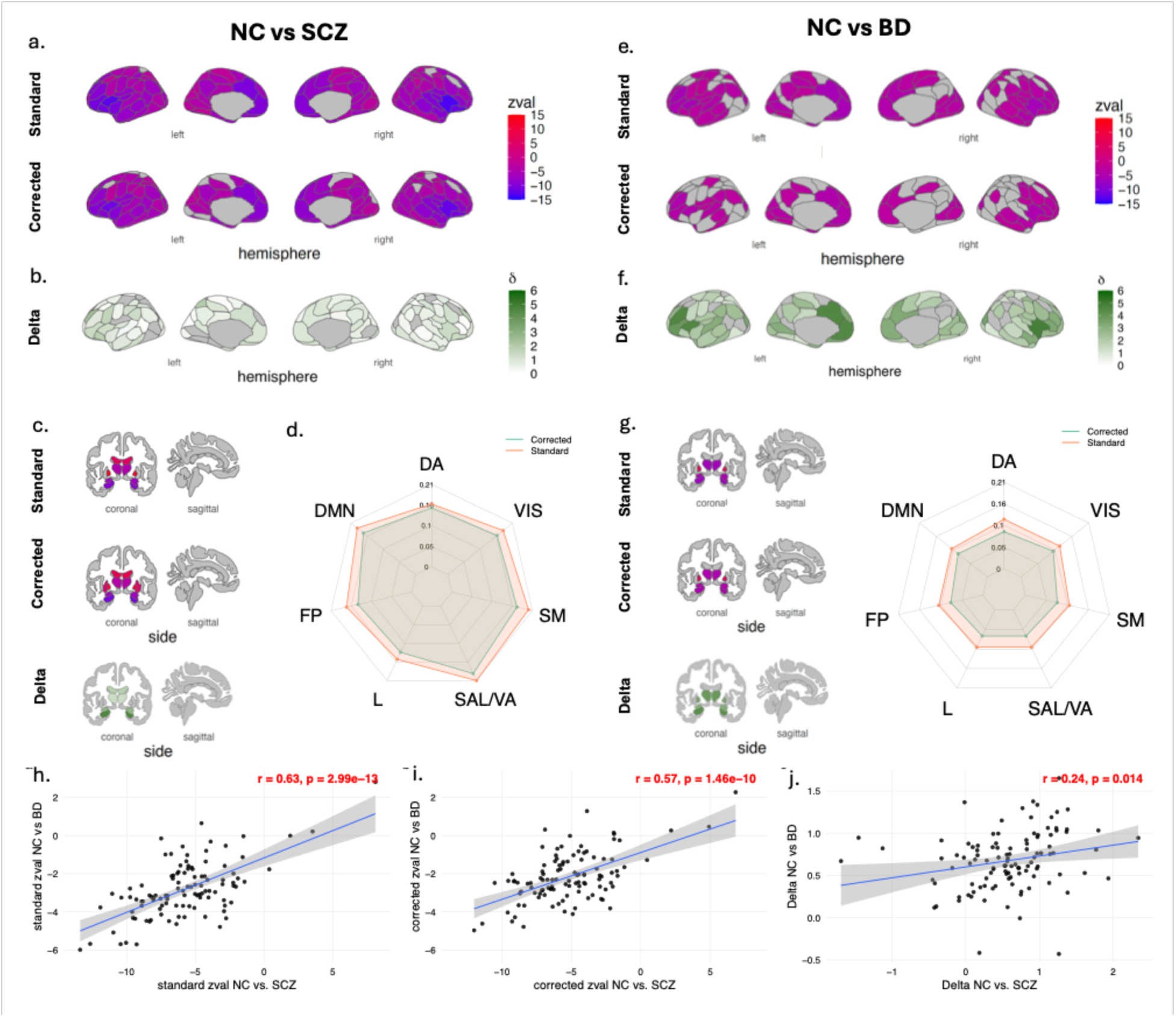
Brain maps show a) the multi-level meta-analysis effects comparing cortical volumes derived from NC vs SCZ and e) NC vs BD across cohorts. Standardized Z-statistics are shown, where negative values correspond to a reduction in grey matter volumes, while positive values correspond to increased grey matter volumes. B-f) Delta values represent the subtraction between Z-statistics of standard and motion-corrected analyses, where higher values correspond to larger effect reductions. C) Meta-analysis effects comparing subcortical volumes from NC vs SCZ and g) NC vs BD across cohorts and related deltas are shown. Results are shown at pFDR<0.05. Radar charts reporting the effects (standardized Z-statistics) of the group effect comparing d) NC vs SCZ and h) NC vs BD for the standard (orange) and motion corrected (green) analyses across seven functional networks. For visualization purposes, Z-statistics were transformed into Cohen’s d for the radar charts to measure effect size. Scatterplots show the relationship between (h) the standardized meta-analytical Z-statistic across cohorts derived from standard analysis, (i) from motion-corrected analysis, and (j) the variation of the effects when correcting for motion parameters, represented by Δs, between SCZ and BD. Pearson’s r and p values are shown. Abbreviations. FP: fronto-parietal network; DMN: default mode network; VIS: visual network; SM: sensorimotor network; SAL/VA: salience and ventral attention network; L: limbic network.

Incorporating motion covariates led to a systematic attenuation of these group differences. Across fMRI scan types and cohorts, 85% of ROIs exhibited a reduction in effect size when motion was accounted for, computed as the difference between standardized estimates derived from standard and motion-corrected models (Δ; Figure 3b). The motion-corrected models yielded 5% fewer ROIs showing a significant group effect (SCZ vs. NC; pFDR<0.05) compared to the standard analysis.

To assess the impact of motion at a larger spatial scale, we aggregated ROIs into seven canonical functional networks and a subcortical network ^38^. At the network level, motion correction consistently reduced the magnitude of SCZ–NC volumetric differences across all networks (Figure 3c), indicating that motion-related effects on estimated group differences are ubiquitous across cortical and subcortical networks. Cohort-specific changes in effect size are shown in Figure S4. To rule out the possibility that the observed differences between standard and motion-corrected analyses arose from reduced degrees of freedom in motion-corrected models (vs. standard models), we conducted permutation analyses in which motion principal components were randomly shuffled 5,000 times. Across cohorts and scan types, standardized group-effect estimates from permuted models were highly similar to those from standard models (r > 0.95; Supplementary Table S4), indicating that attenuation of SCZ–NC effects reflects motion-related characteristics rather than model complexity. Consistent with this interpretation, permutation-based comparisons revealed that in 78% of ROIs, standardized group-effect estimates from motion-corrected models were smaller than those obtained from permuted models (empirical p > 0.5), demonstrating a widespread reduction of apparent SCZ–NC differences attributable to motion correction.

### 2.4 Motion affects group differences comparing BD vs. NC

Consistent with the SCZ-NC findings, accounting for motion led to a systematic attenuation of group differences when comparing patients with BD and NC. Across the CNP, BSNIP, and UNIBA2 cohorts (N = 185 BD patients). The multi-level metanalyses yielded a 24% reduction in the number of ROIs showing significant differences relative to the standard analyses (Figure 3e-g). Regions exhibiting reduced grey matter volume in BD patients compared to NC included the insular cortex, temporoparietal cortices, lateral and ventral prefrontal cortices, hippocampus, amygdala, and thalamus, with increased volume observed in the pallidum. Motion correction resulted in a reduction of effect size in 97% of ROIs, mirroring the pattern observed in SCZ. At the network level, effect sizes were consistently attenuated across all seven cortical networks and the subcortical network (Figure 3h). Cohort-specific results are shown in Figure S5.

To test cross-diagnostic specificity, we quantified the spatial correspondence of effect sizes across ROIs between SCZ vs NC and BD vs NC comparisons using multiple complementary metrics. Among BD-significant ROIs, 100% showed the same direction of effect as in the SCZ comparison, with near-complete concordance also observed across all SCZ-significant ROIs (99%). The Dice coefficient indicated extensive overlap of significant regions (Dice=0.87, with 80 of 108 ROIs significant in both comparisons and only 1 ROI significant exclusively in BD). BD effects accounted for a median of 47% (IQR: 33–65%) of the SCZ effect size magnitude across SCZ-significant ROIs, consistent with attenuated but spatially congruent effects. The Pearson correlation between regional effect sizes was significant in both standard (r=0.63, p=3×10⁻¹³; spin test empirical p=5×10⁻¹⁵; Figure 3h) and motion-corrected analyses (r=0.57, p=1.5×10⁻¹⁰; spin test empirical p=5×10⁻¹⁵; Figure 3i), with motion-related variation of effect sizes (Δ) also showing significant cross-diagnostic association (r=0.24, p=0.014; spin test empirical p=0.006; Figure 3j), and 85% of ROIs showing concordant reduction of effect sizes in both disorders.

To additionally quantify the extent to which motion-related variance overlapped with case–control effects across regions, we computed an ROI-wise estimate of shared variance. We estimated ROI-wise overlap between motion-related effects in NC and case–control effects by combining their partial squared-R values with the spatial correlation between their effect maps. The resulting shared variance was expressed as a fraction of the case–control squared-R, yielding an estimate of the proportion of apparent disease-related variance statistically shared with motion of 6% across regions.

### 2.5 Motion-stratified effects in UK Biobank vs. SCZ-related differences

The large sample size of NC drawn from the UK Biobank (RS: N=5,123; FI: N=4,802) enabled a diagnosis-free test of whether head motion alone produces morphometric patterns resembling those observed in SCZ. Within this cohort, we compared grey matter volumes between NC individuals with high vs. low head motion (top vs. bottom decile of PC1 scores, the component explaining the largest proportion of motion variance) using the same analytic pipeline applied to case-control comparisons (See Methods).

High-motion individuals exhibited widespread reductions in grey matter volume relative to low-motion individuals, with significant differences observed in 79% of ROIs when considering motion PCs derived from FI fMRI and 90% of ROIs when considering motion PCs derived from RS (pFDR<0.05). The spatial distribution of these motion-related reductions closely resembled the SCZ vs. NC morphometric pattern at both cortical and subcortical levels (Figure 4).

**Figure 4.**
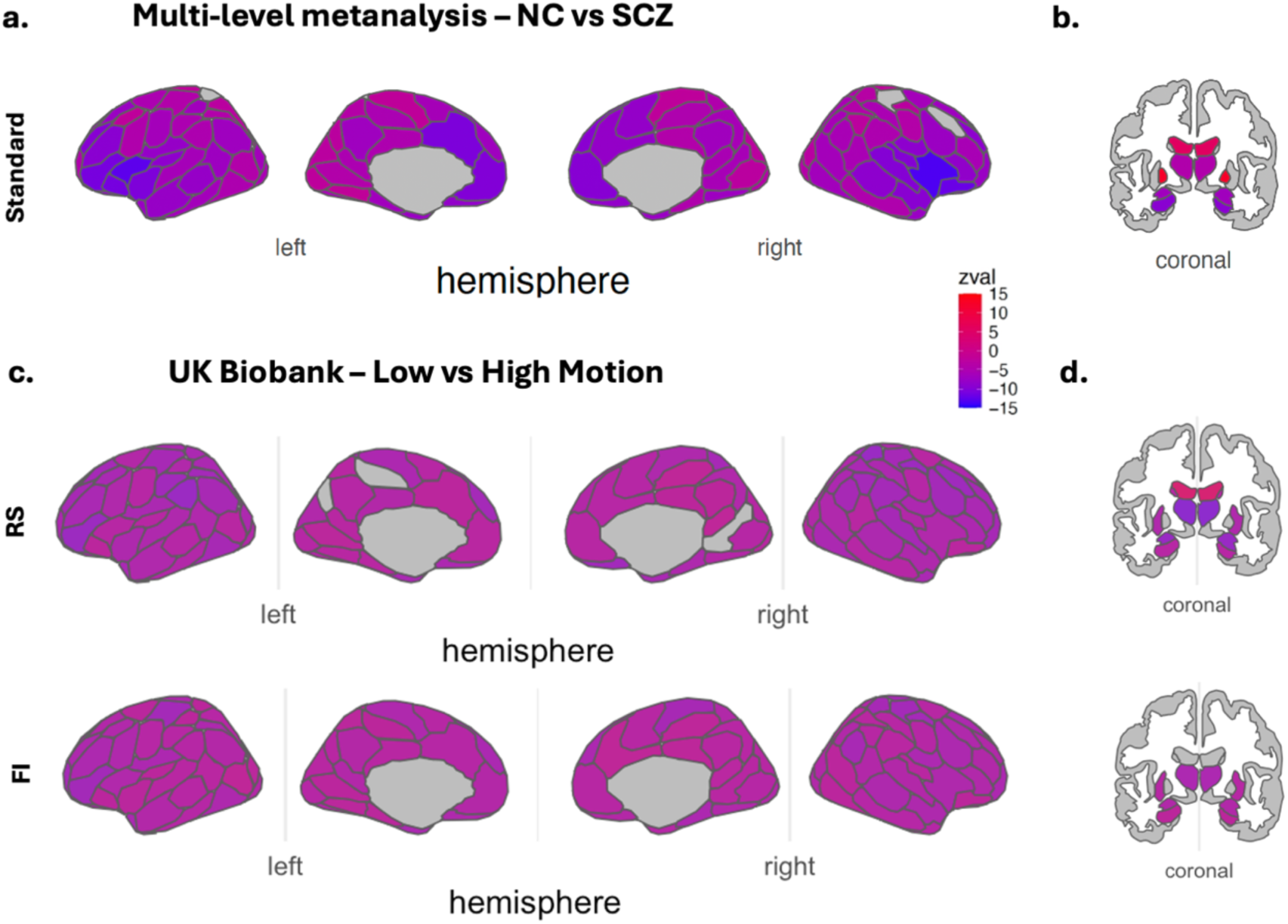
Brain maps show (a-b) the multi-level meta-analysis group effects comparing cortical and subcortical volumes derived from NC vs SCZ (from Figure 3). Standardized Z-statistics are shown, where negative values correspond to a reduction in grey matter volumes, while positive values correspond to increased grey matter volumes. C-d) Group effects comparing cortical and subcortical volumes from low- vs high-motion NC from the UK Biobank. All results are shown at pFDR<0.05.

To quantify this correspondence, we assessed spatial similarity using multiple complementary metrics capturing different aspects of map overlap (see Methods). Among ROIs showing significant SCZ-NC differences, 100% exhibited the same direction of effect in the UKB motion-stratified comparison using RS-derived motion estimates, and 99% using FI-derived estimates. The Dice coefficient indicated extensive overlap between significance maps (RS: Dice=0.91, 91 of 108 ROIs significant in both; FI: Dice=0.84, 76 of 108). To estimate the magnitude of this correspondence, we computed the ratio of absolute z-statistics from the UKB motion-stratified and SCZ-NC comparisons across SCZ-significant ROIs. Motion-stratified differences accounted for a median of 62% (IQR: 42–89%) of the SCZ case-control effect size magnitude using RS-derived estimates, and 45% (IQR: 33–65%) using FI-derived estimates. Spin permutation tests were significant for RS (p=0.003) and for FI (spin test empirical p=0.038). Together, these converging metrics indicate that motion alone, in the absence of any psychiatric diagnosis, is sufficient to generate morphometric differences that reproduce the spatial distribution, direction, and a substantial proportion of the magnitude of SCZ-related grey matter patterns.

## 3. Discussion

Stuctural MRI measurements comparing patients with psychiatric illness and conrol samples have become a foundational standard of clinical research in psychiatry. Differences are almost always reported and results tend to be interpreted as indications of neurodegeneration or atrophy. The fact that MRI is a physico-chemical technology and not strictly an anatomical assay has raised doubts about the underlying causes of the MRI findings^4,5^. We evaluated the impact of estimated in-scanner head motion on morphometric differences between patients with SCZ or BD and NC across eight independent cohorts totaling over 9,000 individuals. By deriving motion estimates from fMRI scans collected within the same session as the structural scan and including these estimates as covariates, we showed that head motion systematically biases case-control morphometric differences, specifically leading to underestimated grey matter. Three primary findings emerged. First, motion explains 1-6% of the variance in grey matter volume estimates in NC, a magnitude comparable to the reported SCZ-related volumetric differences ^21^ and, notably, to reported genetic effects on brain structure ^39,40^. Since genetic influences of this magnitude are considered meaningful in neuroimaging research, motion confounds warrant equal attention. Second, adjusting for motion led to a widespread reduction of group differences, reducing effect sizes in 84% of ROIs in SCZ and 97% in BD analyses, with reductions observed across all cortical and subcortical networks. Third, a falsification analysis in the UK Biobank showed that comparing high-motion versus low-motion NC individuals, in an identical analytical framework as case-control comparisons, recapitulated the spatial pattern of grey matter volume reductions observed in SCZ. In this case, proxies of motion alone, independent of any psychiatric diagnosis, generated morphometric patterns resembling those attributed to illness in distribution and effect size.

### 3.1 Motion effects are replicable across scans

The neuroimaging community is generally aware of the disruptive effect of motion on scanning quality, and also that such an effect goes beyond the generation of readily visible scanning artifacts. Prior work has shown that motion metrics derived from resting-state fMRI account for a non-trivial portion of variance in structural MRI volume estimates ^14^, reflecting a systematic source of bias rather than random noise ^13^. We extended this approach by gathering motion information from multiple scan types, to increase robustness and generalizability of motion characterization. Motion estimates showed consistency across fMRI scan types, with five PCs consistently capturing the dominant axes of variability. More specifically, the first three PCs demonstrated the highest replicability across cohorts and the greatest patient-control differences in both SCZ and BD. These findings extend prior reports in smaller samples ^13^ to a substantially larger sample, indicating that variability in case-control studies partly reflects systematic differences in head motion. Given that motion explained 1–6% of grey matter variance in NC and reported case-control effect sizes in SCZ are of comparable magnitude ^29,37^, these findings underscore its practical relevance for psychiatric neuroimaging. The consistency of motion effects across task-based and resting-state scans supports the broad applicability of this approach.

Within individuals, motion estimates were correlated across different fMRI scans, indicating that participants who moved more during one scan tended to move more across others. This suggests that fMRI-derived motion parameters provide a reasonable proxy for overall in-scanner motion during the session, including during the structural acquisition. Whether motion reflects trait-like characteristics ^20,41^ or is influenced by state-dependent factors, such as session-specific discomfort, remains unresolved. Evidence that head motion shows heritable components ^39,40^ raises the possibility that associations between genetic and morphometric estimates may be partially confounded by shared influences on motion behavior.

### 3.2 Motion affects volumetric differences between patients and controls

The neuroimaging community has long known that motion affects volumetric estimates ^13^, yet this confound has rarely been systematically addressed in large-scale psychiatric case–control studies, despite cautionary notes ^5,14^. Standard analyses here presented replicated established SCZ-related volumetric reductions in the insula, prefrontal cortex, medial parietal regions, and thalamus ^29,37,42–45^ confirming that the samples here analyzed are representative of existing literature. Motion-adjusted analyses systematically reduced these effects at both ROI and network levels, indicating that motion-related bias operates across spatial scales. However, the regional pattern of significant group differences remained largely consistent after correction: key regions including the insula, prefrontal cortex, medial parietal areas, and thalamus retained significant effects, in line with prior literature. Thus, while motion correction attenuated effect sizes, reducing them in 85% of ROIs, the topography of SCZ-related volumetric differences was preserved. These findings suggest that motion, at least as represented in our analyses, inflates rather than fabricates case-control differences, and that established regional findings retain validity, albeit with smaller effect sizes than previously estimated.

The observation that similar motion-related patterns were present in BD analyses, despite less severe clinical manifestations, further suggests that a substantial component of reported morphometric differences may reflect nonspecific factors related to motor behavior rather than diagnosis-specific neurobiology, given that BD patients exhibited yet showed comparable motion-related reductions ^24^.

The UK Biobank analysis provides a crucial interpretive anchor for these findings. Because this cohort comprises psychiatrically and neurologically unaffected individuals, the observation that high-motion NC participants show morphometric reductions overlapping with SCZ-related patterns rules out explanations based on disease biology, medication, or chronic illness. Instead, it demonstrates that motion alone is sufficient to stratify individuals in terms of estimated grey matter differences resembling those attributed to schizophrenia. This within-cohort, diagnosis-free falsification represents a particularly compelling argument in favor of motion-related bias and underscores the risk of attributing such patterns to pathology in the absence of rigorous motion control. It also suggests that the results in the patient-control comparisons may actually underestimate the effects of motion on these differences.

### 3.3 An unresolved debate in psychiatric structural neuroimaging

A common argument of discussion is that motion may reflect meaningful individual differences related to the disorder ^39,40^, thus scrubbing its effect may also remove meaningful disease-related signal. However, this assumption is difficult to reconcile with the lack of consistent postmortem evidence supporting the magnitude and spatial extent of the structural alterations observed in MRI case-control comparisons ^4,24,29^. Because structural MRI reflects physicochemical signal properties rather than direct measurements of tissue composition or cytoarchitecture ^4^, observed differences between patients and controls cannot be assumed to reflect the bona fide neuropathology. Motion is only one of several factors that can influence MRI-derived estimates, alongside medication exposure, substance use, vascular changes, body weight changes, perfusion, cholesterol levels, and other comorbidities ^46^. A conservative and methodologically sound approach is therefore to account for motion explicitly and interpret residual effects with appropriate caution.

We do not propose motion adjustment via fMRI-derived parameters as a definitive solution to motion artifacts in sMRI. Available options to clean structural MRI include prospective motion correction approaches such as PROMO, which use image-based navigators to maintain a fixed measurement coordinate system relative to the head and substantially reduce motion-induced artifacts in high-resolution T1-weighted scans, improving morphometric reliability in both adults and children ^47–50^. In parallel, retrospective correction frameworks operating in k-space or image space, including recent deep-learning–based methods, have shown marked improvements in image quality and cortical surface reconstruction for motion-degraded structural MRI, although these approaches remain computationally demanding and are not yet routinely applicable to large multi-site legacy datasets ^51^. However, reprocessing large legacy datasets from scratch using these approaches remains unfeasible in practice, and large-scale consortium studies are consequently published without any motion correction ^52^. Our approach provides a simple plug-and-play solution that can be readily implemented in any dataset containing concurrent fMRI acquisitions, a feature shared by most contemporary large scale neuroimaging studies. The consistency of effects across multiple scan types highlights the robustness and practical utility of this method, and publicly available scripts accompanying this work enable straightforward integration into existing analysis pipelines when functional data are available (see Data Availability).

### 3.4 Limitations

To maximize inclusion of available data, we analyzed up to three fMRI scans per cohort without explicitly controlling scan order within acquisition protocols. As scan order and task composition can differ between patients and neurotypical controls, unmodeled within-session effects, such as fatigue or habituation, may have contributed additional variability to motion estimates. Moreover, motion parameters were averaged across time points within each scan, yielding summary measures that do not capture temporal fluctuations in movement and therefore represent relatively coarse approximations of motor behavior during scanning.

Our analyses integrated cross-sectional data from multiple cohorts comprising different combinations of resting-state and task-based fMRI sessions, resulting in variation in sample composition and size across scans. Although this heterogeneity may contribute to between-scan variability, it does not affect the primary comparisons between standard and motion-corrected models, which were conducted within matched samples for each analysis. Finally, while inclusion of BD patients enabled an initial assessment of diagnostic generalizability beyond SCZ, extending this framework to additional psychiatric, neurological, and developmental populations will be necessary to fully establish the scope and specificity of motion-related bias in structural MRI.

## 4. Conclusions

Estimated in-scanner head motion substantially correlates with case-control morphometric differences commonly attributed to psychiatric illness. Across multiple cohorts, motion accounted for a meaningful proportion of grey matter variance, attenuated group differences in schizophrenia and bipolar disorder, and, critically, was sufficient to produce SCZ-like morphometric patterns in a large, psychiatrically unaffected sample from the UK Biobank. These findings demonstrate that motion-related bias can inflate and spatially structure neuroanatomical differences found with sMRI. Our findings underscore prior cautions when interpreting MRI data as direct biological measures ^4^ and highlight the need to systematically account for motion in structural analyses. The implications extend beyond psychiatric populations to neurological conditions and lifespan studies where motion differences between groups are expected.

## Data Availability

UK Biobank data are publicly available at http://www.ukbiobank.ac.uk/ (application ID 105731). HCP data are publicly available at https://www.humanconnectome.org/study/hcp-young-adult. CNP data are publicly available at https://openfmri.org/dataset/ds000030/. B-SNIP data are available through the NIH Data Archive at https://nda.nig.gov. LIBD data are available from DRW upon request. DNS data are available from ARH upon request. UNIBA1 and UNIBA2 data cannot be shared at the individual level due to ethical restrictions. Analysis code and data derivatives are publicly available at https://github.com/robertapassiatore/motion-profiling-smri.

## Acknowledgments

We thank Martina Asselti, Dr. Annalisa Lella, Teresa Claudia Pennacchio, and Dr. Alessandra Raio (Department of Translational Biomedicine and Neuroscience, UNIBA) for their help in data collection.

## Funding

European Union MUR PNRR Extended Partnership on Neuroscience and Neuropharmacology, Project PE00000006 CUP H93C22000660006 “MNESYS” (AB, GB, LG, AMM, AR, GP)

Apulian regional government, “Early Identification of Psychosis Risk” (AB)

Ministero della Salute INNOVA, PNC-E3-2022-23683266 PNC-HLS-DA, CUP C43C22001630001 (LAA, AB, GB)

European Union NextGenerationEU, PRIN 2022 PNRR, Prot. P2022HNBJX (GP)

Intramural Research Program of the NIMH, Clinical Brain Disorders Branch, protocol 95-M-0150 (DRW)

## Author contributions

Conceptualization: GP, RP, NS, GS Methodology: RP, NS, GS, AL, CM

Investigation: AL, ALG, ARK, ETW, GB, LF, WSU, Apulian Network on Risk for Psychosis

Formal analysis: RP, NS, GS, AL, CM

Resources: GP, AB, AMM, AR, ARH, DG, DRW, LG

Funding acquisition: LAA, AB, GB, LG, AMM, AR, DRW, GP Supervision: LAA, AB, DRW, GP

Writing -- original draft: RP, NS, GS, GP Writing -- review and editing: all authors

## Supplementary Material

### Methods

#### Samples

We gathered data from the Human Connectome Project – Young Adults (HCP), the Duke Neurogenetics Study (DNS), the UCLA Consortium for Neuropsychiatric Phenomics (CNP), the Bipolar and Schizophrenia Network for Intermediate Phenotypes (BSNIP) consortium, two independent cohorts collected at the University of Bari Aldo Moro using different scanners and protocols (from hereinafter referred as UNIBA1 and UNIBA2), and one provided by the Lieber Institute of Brain Development (LIBD). Additionally, we drew a subset of 5,123 individuals from UK Biobank. Only participants with no evidence of lifetime psychiatric and neurological diagnoses were included, following previously published UK Biobank data selection criteria ^53,54^.

All participants were in good general health and free of conditions known to influence MRI data collection. Inclusion criteria for NC were the absence of any lifetime psychiatric disorder as evaluated with the Structured Clinical Interview for DSM-IV ^55^. Patients with SCZ met criteria for schizophrenia or schizoaffective disorder; patients with BD met criteria for BD type I or II. The Institutional Review Board approved data collection at each institution, and all subjects provided informed consent according to the Declaration of Helsinki. Detailed recruitment, inclusion/exclusion criteria, and protocol descriptions have been published elsewhere ^16,56–59^.

#### Scanning protocols

The scanning protocols comprised a structural and at least one functional scan for all cohorts. Among functional scans, we considered both resting-state and task-based fMRI scans. The resting state scans allowed us to capture inherent motion patterns during rest, while the task-based scans were considered, when available, to detect motion patterns induced by specific task demands, such as button presses.

The resting state was available for all cohorts except UNIBA1. Blocked paradigms of the N-back task ^17,19^ were available for the LIBD, UNIBA1, UNIBA2, and HCP cohorts, which measure increasing spatial working memory loads. An event-related spatial working memory paradigm was available for the CNP cohort ^57^, and a numeric working memory task adapted from Tan et al. ^60,61^ was available for the DNS. For the LIBD, the DNS, and HCP, we also considered a block design implicit emotion recognition task, i.e., the faces matching task ^18^, whereas, for UNIBA1, we used an event-related explicit emotion recognition task based on the identification of the emotions displayed by gender-balanced human faces presenting angry, fearful, happy, and neutral expressions, and an implicit emotion recognition task based on the gender recognition in the same human faces. For UNIBA2, we acquired a revised version of the event-related emotion recognition task used in the UNIBA1 cohort, including only angry, fearful, and neutral facial expressions. The neuropsychological paradigms acquired for each cohort are described elsewhere ^16,57,60,61^.

#### Data acquisition, processing and quality control

Structural MRI acquisition parameters are described in Table 1. All T1-weighted scans were visually inspected to detect artifacts. We estimated gray matter volumes through the Computational Anatomy Toolbox (CAT, https://neuro-jena.github.io/cat/) standalone version 12.8.2 ^62^ for SPM12 (Statistical Parametric Mapping, https://www.fil.ion.ucl.ac.uk/spm/software/spm12/), following the standardized protocol developed for the ENIGMA consortium ^63^. Post-segmentation quality control followed automated procedures ^64^, retaining only segmentations achieving satisfactory quality grades ^65^. Individual grey matter segmentations were normalized to MNI space at 1.5-mm³ and smoothed using a 6-mm FWHM Gaussian kernel.

Functional MRI scans underwent visual quality control and were realigned to the mean volume using SPM12. We did not exclude individuals for excessive motion to preserve variability in motion characteristics. From the six automatically extracted realignment parameters, we derived 18 motion measures ^7,8,25,66^ detailed below.

### Data Analysis

#### Principal Component Analysis

We extracted eighteen motion parameters for each fMRI scan collected for the eight cohorts. The eighteen motion parameters included six rotation parameters representing mean and maximum values on the x, y, and z axes; six translation parameters also covering mean and maximum values on these axes; four parameters measured Euclidean distance for voxel shifts in MRI space ^8^, and two assessed framewise displacement, indicating movement between images ^66^. Motion parameters were preliminary transformed to mitigate deviation from the Gaussian distribution using a natural-log function, standardized across the entire sample.

To reduce the number of predictors, thereby mitigating overfitting and enhancing model generalization and replicability, we performed a PCA on the log-transformed eighteen motion parameters Direct *oblimin* rotation was used, permitting correlation between components.

#### ROI-based analyses

Grey matter volume estimates were extracted from 100 bilateral cortical regions using the Schaefer-Yeo atlas ^28^ and 8 subcortical structures (hippocampus, amygdala, thalamus, nucleus accumbens, caudate, putamen, pallidum, lateral ventricles) from the Hammersmith atlas ^30^, selected based on ENIGMA Schizophrenia Working Group findings ^29^ and relevance to psychiatric^65^ and neurologic conditions ^67^. Bilateral volumes were computed as the sum of hemispheres ^29^. Scanner effects were harmonized using neuroCombat ^68^.

The variance explained by the 5 motion PCs was tested in NC using linear regression, controlling for age, age-squared, sex, and TIV. For case-control analyses (SCZ vs. NC; BD vs. NC), we compared standard models (covariates: age, age-squared, sex, TIV) with motion-corrected models that additionally included the 5 motion PCs. Analyses were conducted separately for each cohort and fMRI scan type.

To generalize findings, standardized statistics were entered into multi-level meta-analyses using the *rma.mv* function in R metafor ^69^, with random intercepts for cohorts and sessions. FDR correction was applied across 108 ROIs. The impact of motion correction was quantified as the percentage reduction in significant results:

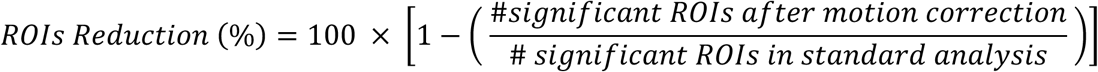

Effect size differences were calculated as the delta (Δ) between absolute Z-statistics from standard and motion-corrected analyses. Results were summarized by aggregating cortical ROIs into 7 networks plus a subcortical network (Figure 3). Spatial correspondence between SCZ and BD effects was tested using spin permutation tests ^70,71^ with 10,000 rotations.

Variance Inflation Factor values (range: 1–1.2) indicated no multicollinearity concerns. Permutation analyses (5,000 iterations) confirmed that observed differences between standard and motion-corrected analyses were not attributable to differences in degrees of freedom.

#### UK Biobank motion-stratified analyses

To test whether in-scanner head motion alone is sufficient to reproduce morphometric patterns resembling those observed in SCZ, we conducted a falsification analysis in the UKB sample of psychiatrically and neurologically unaffected participants. From the initial samples with available structural MRI, RS (N=5,123) and/or FI (N=4,802) fMRI data, participants were ranked by PC1 motion scores, the component explaining the largest proportion of motion variance, and stratified into high-motion (top 10%) vs. low-motion (bottom 10%) groups. This procedure yielded two comparably sized groups for RS (high-motion: N=521, age=53±7, 54% male; low-motion: N=518, age=51±7, 40% male) and for FI (high-motion: N=482, age=55±8, 40% male; low-motion: N=481, age=50±8, 59% male). Regional grey matter volume differences were assessed using linear models controlling for age, age-squared, sex, and TIV, mirroring the case-control analysis and correcting for multiple comparisons using FDR. Spatial similarity between high- vs low-motion effects and SCZ vs NC effects was quantified using spin permutation tests ^70,71^ 10,000 rotations) on ROI-wise z-scores across all cortical and subcortical regions without statistical thresholding.

Spatial similarity between the UKB motion-stratified effects and SCZ-NC effects was assessed using multiple complementary metrics. Pearson correlation and spin permutation tests (10,000 rotations) were applied to ROI-wise z-statistics without statistical thresholding. The Dice coefficient was computed to quantify the overlap of statistically significant ROIs (pFDR<0.05) between the two maps, defined as Dice = 2|A∩B| / (|A|+|B|), where A and B denote the sets of significant ROIs in the SCZ-NC and UKB motion-stratified analyses, respectively. Directional concordance was defined as the proportion of SCZ-significant ROIs that showed the same sign of the z-statistic in the UKB comparison. To estimate the magnitude of overlap, we computed the ratio of absolute z-statistics (|z_UKB| / |z_SCZ|) for each SCZ-significant ROI and summarized these ratios using the median and interquartile range. This ratio quantifies the proportion of the SCZ case-control effect size that motion alone can reproduce in a diagnosis-free sample, independently of the linear association between the two maps.

## Supplementary tables

**Table S1.** Cross-modality correlations of the five motion-PCs within each cohort. The blank quadrants indicate the lack of overlap between participants included in the analyses across different fMRI scans. *Abbreviations*. WM: Working memory; FE: Faces – Explicit; FI: Faces – Implicit; RS: Resting-State. Pearson’s r and two-sided p-value are reported: *** p < 0.001, ** p < 0.01, * p < 0.05

| RS-FI fMRI | PC1 | PC2 | PC3 | PC4 | PC5 |
| --- | --- | --- | --- | --- | --- |
| UK Biobank | R = 0.27 *** | R = 0.16 *** | R = 0.13 *** | R = 0.14 *** | R = 0.10 *** |
| LIBD | R = 0.26 *** | R = 0.13 * | R = 0.08 ns | R = 0.08 ns | R = 0.03 ns |
| UNIBA2 | R = 0.42 *** | R = 0.49 *** | R = 0.26 *** | R = 0.30 *** | R = 0.40 *** |
| DNS | R = 0.42 *** | R = 0.30 *** | R = 0.25 *** | R = 0.27 *** | R = 0.41 *** |
| HCP | R = 0.24 *** | R = 0.63 *** | R = 0.09 ** | R = 0.04 ns | R = 0.01 ns |
| WM-FI fMRI | PC1 | PC2 | PC3 | PC4 | PC5 |
| LIBD | R = 0.52 *** | R = 0.19 *** | R = 0.10 ns | R = 0.11 * | R = 0.19 *** |
| UNIBA1 | R = 0.30 *** | R = 0.10 ns | R = 0.23 *** | R = 0.10 ns | R = 0.10 ns |
| UNIBA2 | R = 0.41 *** | R = 0.52 *** | R = 0.23 * | R = 0.35 ** | R = 0.08 ns |
| DNS | R = 0.35 *** | R = 0.31 *** | R = 0.44 *** | R = 0.28 *** | R = 0.37 *** |
| HCP | R = 0.29 *** | R = 0.68 *** | R = 0.20 *** | R = 0.09 ** | R = 0.09 ** |
| WM-RS fMRI | PC1 | PC2 | PC3 | PC4 | PC5 |
| LIBD | R = 0.22 *** | R = 0.08 ns | R = 0.17 ** | R = 0.05 ns | R = 0.08 ns |
| UNIBA2 | **R = 0.23 \*\*\*** | **R = 0.69 \*\*\*** | **R = 0.29 \*\*\*** | **R = 0.28 \*\*\*** | **R = 0.16 \*\*\*** |
| CNP | **R = 0.81 \*\*\*** | **R = 0.34 \*\*\*** | **R = 0.33 \*\*\*** | **R = 0.39 \*\*\*** | R = 0.13 ns |
| HCP | **R = 0.29 \*\*\*** | **R = 0.68 \*\*\*** | **R = 0.20 \*\*\*** | **R = 0.09 \*\*** | **R = 0.09 \*\*** |
| FE-FI fMRI | PC1 | PC2 | PC3 | PC4 | PC5 |
| UNIBA1 | **R = 0.52 \*\*\*** | **R = 0.43 \*\*\*** | **R = 0.49 \*\*\*** | **R = 0.21 \*\*** | R = 0.10 ns |
| FE-WM fMRI | PC1 | PC2 | PC3 | PC4 | PC5 |
| UNIBA1 | **R = 0.45 \*\*\*** | **R = 0.16 \*** | **R = 0.28 \*\*\*** | **R = 0.26 \*\*\*** | R = 0.03 ns |

**Table S2.** Statistical results comparing motion-derived PCs in patients with SCZ compared with NC. T-statistics, degrees of freedom, and p-values are reported. Significant differences (p<0.05) are marked in bold font.

| RS fMRI | PC1 | PC2 | PC3 | PC4 | PC5 |
| --- | --- | --- | --- | --- | --- |
| B-SNIP | **t = 5.5795, df = 261.54, p-value = 6.02e-08** | t = 1.8792, df = 286.98, p-value = 0.06123 | t = 1.3257, df = 288.1, p-value = 0.186 | t = 1.3476, df = 295.79, p-value = 0.1788 | t = -0.023239, df = 307.97, p-value = 0.9815 |
| LIBD | **t = 2.0761, df = 221.92, p-value = 0.03904** | **t = 4.2521, df = 191.46, p-value = 3.307e-05** | **t = 2.365, df = 194.45, p-value = 0.01902** | t = 1.4884, df = 205.62, p-value = 0.1382 | t = 1.8107, df = 236.55, p-value = 0.07146 |
| UNIBA2 | **t = 4.2718, df = 65.126, p-value = 6.447e-05** | **t = 4.8034, df = 51.957, p-value = 1.367e-05** | **t = 0.1963, df = 58.656, p-value = 0.8451** | t = 0.51575, df = 59.994, p-value = 0.6079 | t = 1.0602, df = 65.414, p-value = 0.293 |
| **CNP** | **t = 4.0175, df = 81.173, p-value = 0.0001307** | t = 0.26873, df = 76.871, p-value = 0.7889 | t = 0.68026, df = 93.87, p-value = 0.498 | t = 0.90388, df = 66.932, p-value = 0.3693 | t = -1.7522, df = 82.866, p-value = 0.08345 |
| **WM fMRI** | **PC1** | **PC2** | **PC3** | **PC4** | **PC5** |
| **LIBD** | **t = 3.5813, df = 177.42, p-value = 0.0004413** | t = 0.52907, df = 199.15, p-value = 0.5973 | t = 0.093937, df = 203.5, p-value = 0.9253 | t = -0.053833, df = 216.48, p-value = 0.9571 | t = -0.21644, df = 208.16, p-value = 0.8289 |
| **UNIBA1** | **t = 5.7323, df = 71.915, p-value = 2.163e-07** | **t = 3.0831, df = 74.922, p-value = 0.002867** | **t = 2.1374, df = 84.284, p-value = 0.03546** | t = 1.5116, df = 82.18, p-value = 0.1345 | **t = 2.3907, df = 78.485, p-value = 0.01921** |
| **UNIBA2** | **t = 2.1842, df = 52.049, p-value = 0.03348** | **t = 4.8912, df = 50.239, p-value = 1.072e-05** | t = 1.5306, df = 60.853, p-value = 0.1311 | t = 0.84461, df = 61.154, p-value = 0.4016 | t = 0.57772, df = 65.959, p-value = 0.5654 |
| **CNP** | **t = 5.3411, df = 79.794, p-value = 8.5e-07** | t = 0.73464, df = 77.531, p-value = 0.4648 | t = 0.66997, df = 68.802, p-value = 0.5051 | t = -0.86224, df = 72.804, p-value = 0.3914 | t = -0.52206, df = 89.738, p-value = 0.6029 |
| **FI fMRI** | **PC1** | **PC2** | **PC3** | **PC4** | **PC5** |
| **LIBD** | **t = 3.7346, df = 151.71, p-value = 0.0002655** | t = 1.7106, df = 164.89, p-value = 0.08904 | t = 0.92784, df = 171.54, p-value = 0.3548 | t = 0.43394, df = 156.07, p-value = 0.6649 | t = -0.10282, df = 195.98, p-value = 0.9182 |
| **UNIBA1** | t = 1.3092, df = 47.733, p-value = 0.1967 | **t = 4.7166, df = 45.33, p-value = 2.318e-05** | t = 1.719, df = 44.174, p-value = 0.09261 | **t = 2.467, df = 45.101, p-value = 0.01749** | t = 1.6722, df = 44.977, p-value = 0.1014 |
| **UNIBA2** | **t = 2.3945, df = 44.777, p-value = 0.02089** | **t = 3.8621, df = 38.608, p-value = 0.0004171** | t = 1.3754, df = 43.975, p-value = 0.176 | **t = 2.2765, df = 45.078, p-value = 0.02762** | t = -0.36246, df = 44.557, p-value = 0.7187 |
| **FE fMRI** | **PC1** | **PC2** | **PC3** | **PC4** | **PC5** |
| **UNIBA1** | **t = 6.0231, df = 72.227, p-value = 6.532e-08** | **t = 2.0425, df = 66.671, p-value = 0.04506** | **t = 3.6708, df = 74.344, p-value = 0.0004525** | **t = 2.5396, df = 66.585, p-value = 0.01344** | t = 1.9148, df = 72.75, p-value = 0.05945 |

**Table S3.** Statistical results comparing motion-derived PCs in patients with BD compared with NC. T-statistics, degrees of freedom, and p-values are reported. Significant differences (p<0.05) are marked in bold font.

| RS fMRI | PC1 | PC2 | PC3 | PC4 | PC5 |
| --- | --- | --- | --- | --- | --- |
| B-SNIP | **t = 3.4925, df = 142.3, p-value = 0.0006378** | t = 0.45129, df = 164.92, p-value = 0.6524 | t = 0.15079, df = 172.78, p-value = 0.8803 | t = 0.59715, df = 162.51, p-value = 0.5512 | t = -0.12075, df = 192.61, p-value = 0.904 |
| UNIBA2 | **t = 2.259, df = 62.403, p-value = 0.02738** | **t = 4.0733, df = 53.978, p-value = 0.0001528** | **t = 2.7978, df = 63.085, p-value = 0.006816** | t = 0.59229, df = 65.332, p-value = 0.5557 | t = 0.70747, df = 63.446, p-value = 0.4819 |
| CNP | **t = 2.4296, df = 84.586, p-value = 0.01723** | t = 0.2019, df = 89.417, p-value = 0.8405 | t = 0.2093, df = 96.391, p-value = 0.8347 | t = 0.061082, df = 84.261, p-value = 0.9514 | t = 0.12677, df = 85.146, p-value = 0.8994 |
| WM fMRI | PC1 | PC2 | PC3 | PC4 | PC5 |
| UNIBA2 | **t = 2.1659, df = 49.965, p-value = 0.03512** | **t = 4.4371, df = 46.08, p-value = 5.635e-05** | t = 1.471, df = 53.065, p-value = 0.1472 | t = 0.6745, df = 54.424, p-value = 0.5029 | t = 0.41302, df = 61.103, p-value = 0.681 |
| CNP | **t = 3.2469, df = 75.594, p-value = 0.00174** | t = 1.8614, df = 87.545, p-value = 0.06604 | t = 0.3578, df = 86.382, p-value = 0.7214 | t = 0.33897, df = 78.342, p-value = 0.7355 | t = 0.80801, df = 89.727, p-value = 0.4212 |
| FI fMRI | PC1 | PC2 | PC3 | PC4 | PC5 |
| UNIBA2 | **t = 3.1253, df = 73.603, p-value = 0.002542** | **t = 3.7978, df = 55.097, p-value = 0.0003661** | t = 1.3754, df = 43.975, p-value = 0.176 | **t = 2.6223, df = 60.265, p-value = 0.01104** | **t = 2.4477, df = 66.953, p-value = 0.01701** |

**Table S4.** Pearson’s r between the average standardized beta coefficient of the group effect across permutations (N=5,000) and effect sizes obtained in the standard analyses. All p-values are <0.05.

| **Resting state fMRI** | **Pearson's R SCZ** | **Pearson's R BD** |
| --- | --- | --- |
| **B-SNIP** | 0.96 | 0.98 |
| **LIBD** | 0.99 | - |
| **UNIBA2** | 0.99 | 0.98 |
| **CNP** | 0.95 | 0.99 |
| **Working memory task fMRI** | **Pearson's R SCZ** | **Pearson's R BD** |
| **LIBD** | 0.99 | - |
| **UNIBA1** | 0.99 | - |
| **UNIBA2** | 0.98 | 0.99 |
| **CNP** | 0.95 | 0.98 |
| **Implicit emotion recognition task fMRI** | **Pearson's R SCZ** | **Pearson's R BD** |
| **LIBD** | 0.99 | - |
| **UNIBA1** | 0.99 | - |
| **UNIBA2** | 0.97 | 0.98 |
| **Explicit emotion recognition task fMRI** | **Pearson's R SCZ** | **Pearson's R BD** |
| **UNIBA1** | 0.99 | - |

## Supplementary Figures

**Figure S1.**
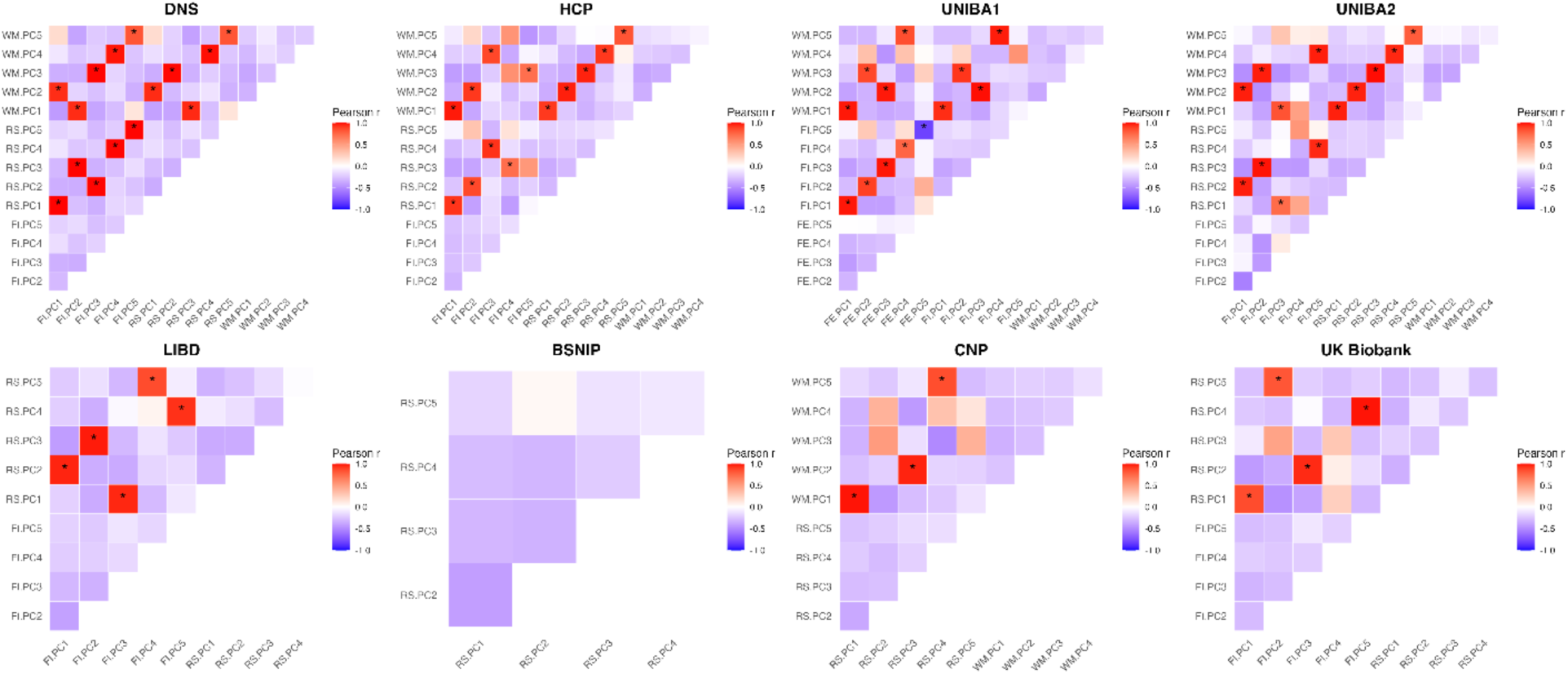
Correlation (Pearson’s r) between the loadings of the eighteen motion parameters across the different cohorts and fMRI scans, with significant associations (pFDR < .05) indicated by a star. *Abbreviations*. WM: Working memory; FE: Faces – Explicit; FI: Faces – Implicit; RS: Resting-State.

**Figure S2.**
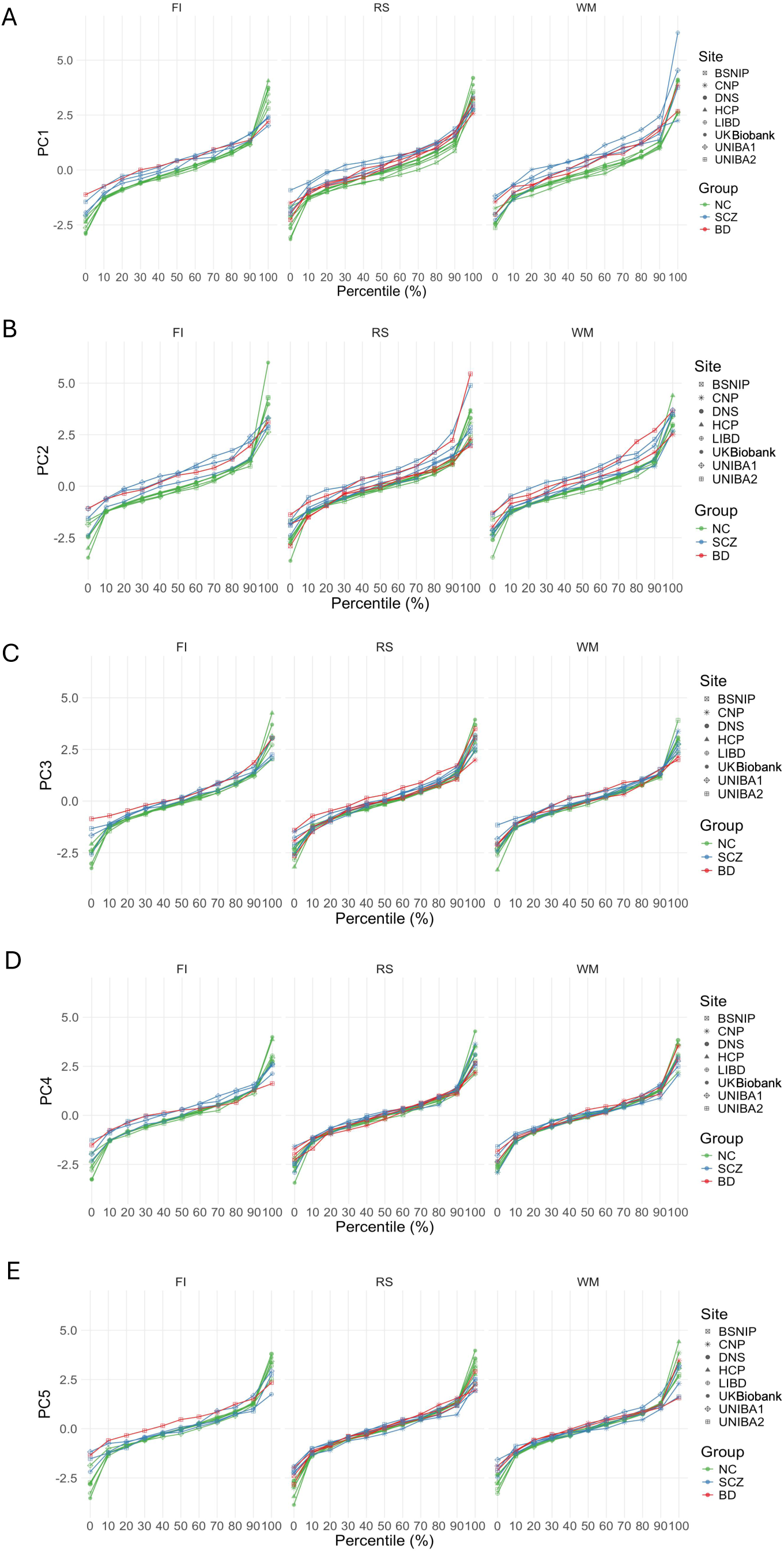
Percentile distribution of motion PCs across cohorts and fMRI scans. *Abbreviations*. WM: Working Memory; FE: Faces – Explicit; FI: Faces – Implicit; RS: Resting-State.

**Figure S3.**
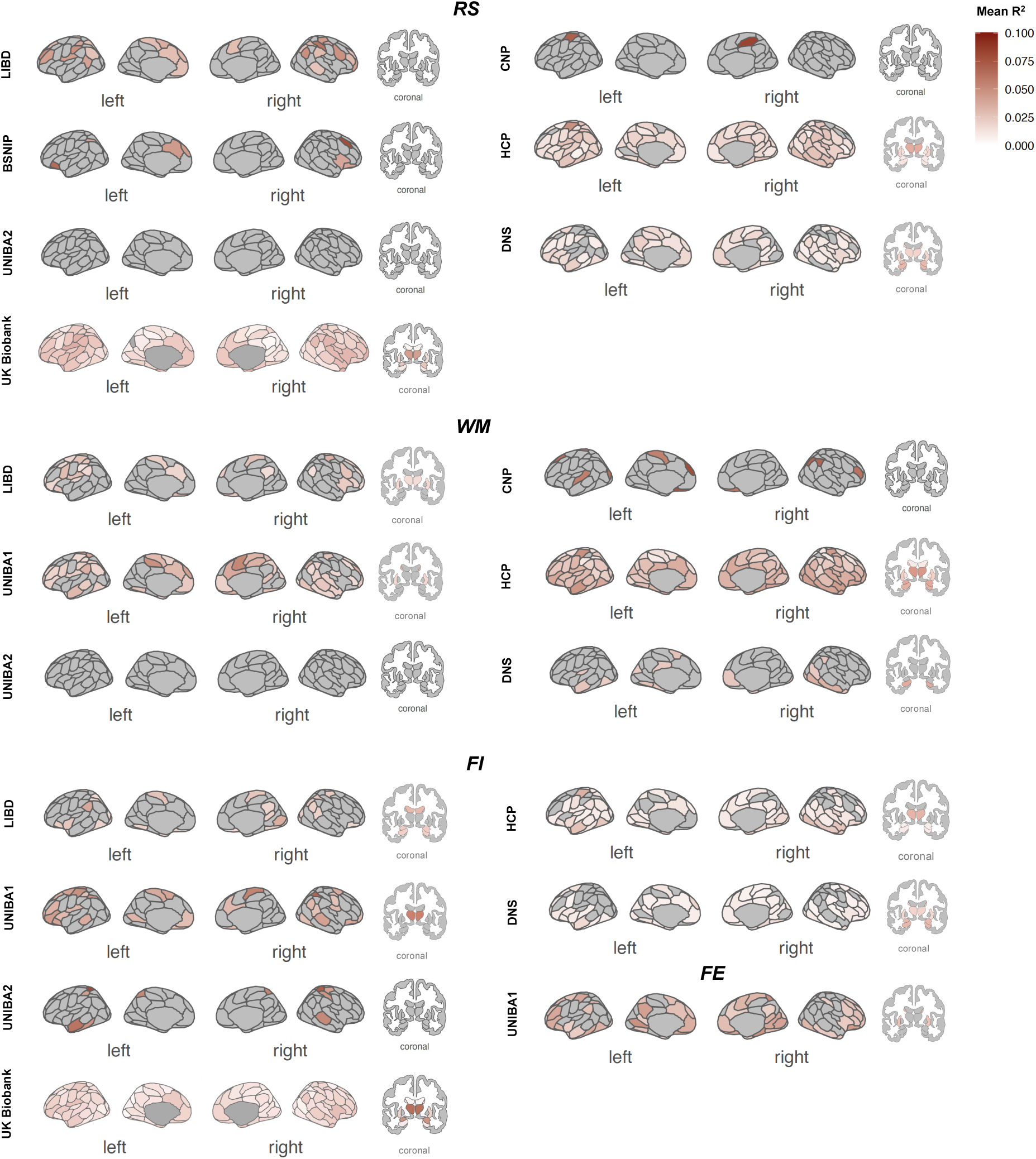
Glass brains show the adjusted R-squared obtained from the full model effects of 5 motion-PCs on grey matter volume estimated in cortical and subcortical ROIs for each cohort and fMRI session. Results are shown at p<0.05. *Abbreviations*. WM: Working Memory; FE: Faces – Explicit; FI: Faces – Implicit; RS: Resting-State.

**Figure S4.**
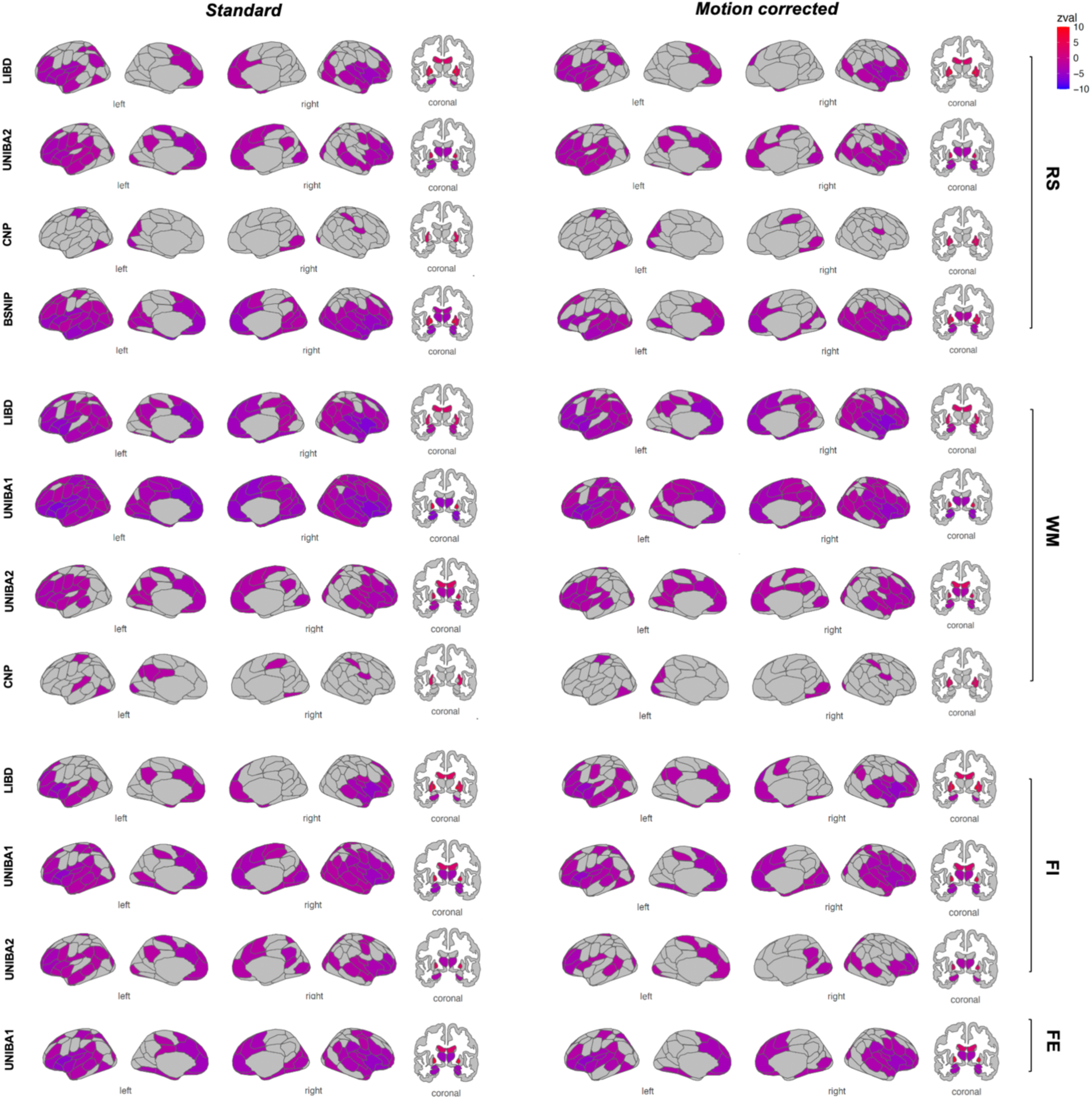
Glass brains show the standardized effects obtained from the standard and motion-corrected analyses comparing grey matter volume in NC vs SCZ for each cohort and fMRI session. Results are shown at p<0.05. *Abbreviations*. WM: Working Memory; FE: Faces – Explicit; FI: Faces – Implicit; RS: Resting-State.

**Figure S5.**
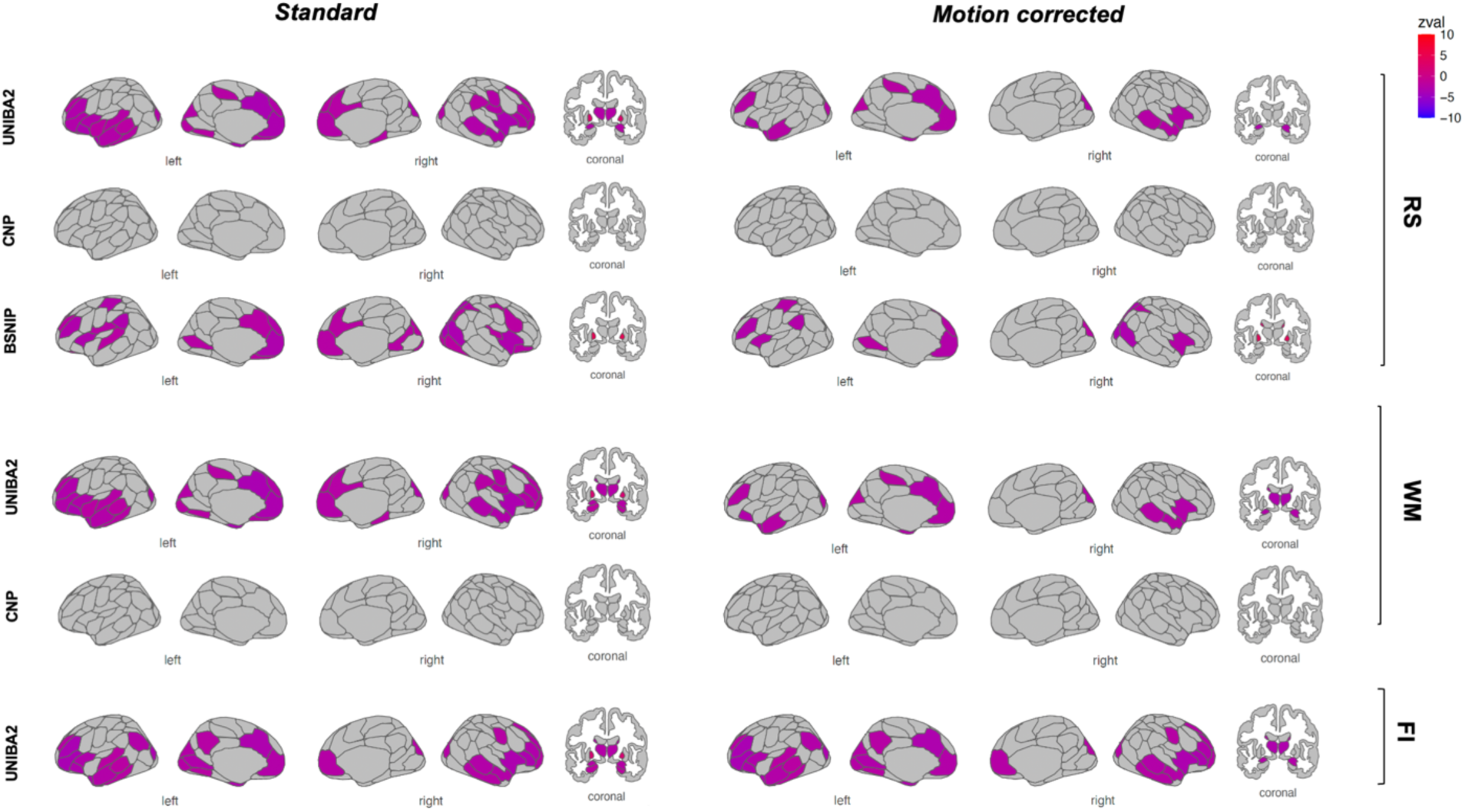
Glass brains show the standardized effects obtained from the standard and motion-corrected analyses comparing grey matter volume in NC vs BD for each cohort and fMRI session. Results are shown at p<0.05. *Abbreviations*. WM: Working Memory; FE: Faces – Explicit; FI: Faces – Implicit; RS: Resting-State.

